# Assessment of COVID-19 vaccine hesitancy among Zimbabweans: A rapid national survey

**DOI:** 10.1101/2021.06.24.21259505

**Authors:** Paddington T. Mundagowa, Samantha N. Tozivepi, Edward T. Chiyaka, Fadzai Mukora-Mutseyekwa, Richard Makurumidze

**Affiliations:** Africa University, College of Health, Agriculture & Natural Science, Clinical Research Centre; Kent State University, College of Public Health, Ohio, USA; University of Zimbabwe, Faculty of Medicine and Health Sciences, Public and Global Health Unit, Harare, Zimbabwe; Institute of Tropical Medicine, Clinical Sciences Department, Antwerp, Belgium; Free University of Brussels (VUB), Gerontology, Faculty of Medicine & Pharmacy, Brussels, Belgium; Zimbabwe College of Public Health Physicians, Harare, Zimbabwe

**Keywords:** COVID-19, coronavirus, vaccine, hesitancy, willingness, Zimbabwe

## Abstract

**Background:** To minimise the devastating effects of the coronavirus disease 2019 (COVID-19) pandemic, scientists hastily developed a vaccine. However, the scale-up of the vaccine is likely to be hindered by the widespread social media misinformation. We, therefore, conducted a study to assess the COVID-19 vaccine hesitancy among Zimbabweans.

**Methods:** We conducted a descriptive online cross-sectional survey using a self-administered questionnaire among adults. The questionnaire assessed willingness to be vaccinated; socio-demographic characteristics, individual attitudes and perceptions, effectiveness, and safety of the vaccine. Multivariable logistic regression analysis was utilized to examine the independent factors associated with vaccine uptake.

**Results:** We analysed data for 1168 participants, age range of 19-89 years with the majority being females (57.5%). Half (49.9%) of the participants reported that they would accept the COVID-19 vaccine. The majority were uncertain about the effectiveness of the vaccine (76.0%) and its safety (55.0%). About half lacked trust in the government’s ability to ensure the availability of an effective vaccine and 61.0% mentioned that they would seek advice from a healthcare worker to vaccinate. Age 55 years and above [vs 18-25 years - Adjusted Odds Ratio (AOR): 2.04, 95% Confidence Interval (CI): 1.07-3.87], chronic disease [vs no chronic disease - AOR: 1.72, 95%CI: 1.32-2.25], males [vs females - AOR: 1.84, 95%CI: 1.44-2.36] and being a healthcare worker [vs not being a health worker – AOR: 1.73, 95%CI: 1.34-2.24] were associated with increased likelihood to vaccinate. History of COVID-19 infection [vs no history - AOR: 0.45, 95%CI: 0.25-0.81) and rural residence [vs urban - AOR: 0.64, 95%CI: 0.40-1.01] were associated with reduced likelihood to vaccinate.

**Conclusion:** We found half of the participants willing to vaccinate against COVID-19. The majority lacked trust in the government and were uncertain about vaccine effectiveness and safety. The policymakers should consider targeting geographical and demographic groups which were unlikely to vaccinate with vaccine information, education, and communication to improve uptake.

## Introduction

Since the first case of the coronavirus disease (COVID-19) caused by the novel severe acute respiratory syndrome coronavirus 2 (SARS-CoV-2) was reported in 2019, more than three million fatalities and 1.55 billion cases have been recorded globally (1) The healthcare systems have been strained and the adverse socio-economic and psychological impacts are overwhelming (2–4).

Fortunately, more than 100 vaccines have gone beyond the pre-clinical development phase with more than half of these reaching the clinical development phase (5). The herd immunity for SARS-CoV-19 can be reached by vaccinating about 60-72% of the population (6), thus, vaccine acceptance rates will play a major role in combating the pandemic. However, vaccine hesitancy has been recognized as a major threat to the control of vaccine preventable diseases (7). The recent COVID-19 pandemic has caused an outbreak of “infodemics” which has led to rapid and far-reaching spread of inaccurate information on the COVID-19 vaccine (8). This deluge of unreliable information can contribute to COVID-19 vaccine hesitancy despite the availability of safe and effective vaccines (9).

Like many other developing countries, Zimbabwe has started the process of securing COVID-19 vaccine. A COVID-19 national deployment and vaccination strategy (10) has been developed and plans for national deployment and training of healthcare workers are underway. In this barrage of infodemics, Zimbabweans have not been spared as conspiracy beliefs filtered through the population, particularly through the different social media platforms. The best way to fight misinformation is through aggressive dissemination of accurate information about the truths of the risks and benefits of COVID-19 vaccine.

Evidence on the population’s intention to be vaccinated for COVID-19 is limited and public health authorities are confronted with a challenge of objectively discerning the truth from the circulating information blast. Understanding the factors influencing vaccine hesitancy is pertinent in crafting targeted communication and interventions to sell the idea of vaccination to specific groups of people. We therefore conducted a national survey to assess COVID-19 vaccine hesitancy among Zimbabweans. We assessed socio-demographic characteristics, individual attitudes and perceptions, Covid-19 vaccine effectiveness and safety, and risks of contracting the COVID-19 in relation to the uptake of the COVID-19 vaccine.

## Methods

### Study Design

We conducted an online descriptive cross-sectional survey in February 2021.

### Study setting

The study was conducted in Zimbabwe. By the end of 2020, the country had an estimated population of 15.1 million (11). Approximately 99.6% of the population is of African origin and the median age is 18.7 years (12). Geographically, the country is divided into 10 provinces and 63 districts. The first COVID-19 case was recorded on the 20^th^ of March 2020 and according to the Zimbabwean Ministry of Health and Child Care Daily COVID-19 Update Report of the 11^th^ of May 2021, the country had recorded a cumulative of 38 466 cases, 36 277 recoveries, and 1579 deaths (13).

### Study participants

All Zimbabwean residents were eligible to participate in the survey. Considering the online nature of the survey and the easy sharing via social networks, we allowed Zimbabweans out of the country to participate but were excluded from the analysis because (information to inform local context) their COVID-19 vaccine uptake was more likely to be influenced by their current context. We included adults aged 18 years and above and those who had participated but below 18 years were excluded. The survey was disseminated mainly via social media networks (WhatsApp and Facebook) hence those registered in those platforms were likely to participate.

#### Sample size

The minimum sample size was estimated at 423, based on the following assumptions; 95% confidence interval, 5% margin of error, the expected proportion of vaccine uptake estimated as 52% (14) in the adult population in Zimbabwe and the attrition rate of 10%.

### Data collection tools and procedures

A multi-item survey questionnaire was developed based on literature review (15,16). The online questionnaire was designed using Google Forms. The survey questionnaire was developed in English and later translated into the two main local languages in Zimbabwe, Shona and Ndebele. The survey questionnaire comprised of five sections with the first section collecting participant’s demographic data (age, sex, residence, educational level, employment status, medical aid status). The second section consisted of questions soliciting information and knowledge on COVID-19 vaccine. The third section sought to assess vaccine uptake, effectiveness and safety. The fourth section assessed the risk of contracting COVID-19. We also collected qualitative information through open-ended questions and will be reported separately.

Our outcome variable was vaccine uptake in the event that a COVID-19 vaccine becomes available and it had three responses, “Would Accept”, “Unsure” and “Would reject”. However, for our analysis we combined our responses into binary those that will be vaccinated “Yes” (Would Accept) and those that would not be vaccinated, “No” (Unsure/Would reject). Potential study participants were recruited using random sampling via social media mainly WhatsApp and Facebook through the network and contacts of the researchers. The researchers encouraged participants to share the online survey with others in their networks as well. The survey was also shared in social media groups for academic and community organizations. Measures were put in place to ensure that the questionnaire was taken only once. Data were collected over a period of 15 days from the 3^rd^ of February 2021 to the 17^th^ of February 2021.

### Statistical Analysis

A database was created in Microsoft Excel using the data extracted from the online server. Responses from the Shona and Ndebele questionnaires were coded into English and then merged into one English database. Data cleaning was done in Excel before exporting it to SPSS for analyses. Statistical analysis was performed using SPSS (V.26.0). Descriptive statistics were conducted using frequencies and proportions and were presented in tables and graphs. Logistic regression models were used to examine factors associated with vaccine uptake. Statistical significance was set at p< 0.05 and 95% confidence interval.

### Ethical considerations

Ethical clearance to carry out this study was obtained from the Medical Research Council of Zimbabwe (MRCZ/A/2714). Informed consent was provided online and all participants were asked to accept or reject participation in the online survey. Participation in this study was voluntary and was not incentivized. All the responses provided during this survey were anonymous.

## Results

### Study participants

We received 1290 responses during the survey period. In terms of language, the distribution of responses was as follows: 1196 English, 71 Shona and 23 Ndebele. Of the 1290 participants who completed the survey, 122 were excluded from analysis for the following reasons (89 were based outside Zimbabwe, 4 were under the age of 18 years, 26 were duplicate responses, and in 3 age was missing). We finally analysed 1168 participants. The study participants were from all the 10 provinces of Zimbabwe and half (50.5%) were from Harare province which houses the capital city of the country.

#### Socio-demographic characteristics of the study participants

A total of 1168 participants were included in the study with a median age of 39 years (interquartile range; IQR: 32-49) and an age range of 19 – 89 years. Table 1 shows a summary of the socio-demographic characteristics of the participants. The majority of the study participants were in the 36-45 years age group. Females constituted (57.5%) while 36% were health care workers. A significantly higher proportion (92.6%) of participants was residing in an urban setting. About 41.2% of the participants reported having at least one chronic condition. More than 90% reported having a good health status and a similar proportion had tertiary education as the highest level of education achieved. About 5% reported having tested positive for COVID-19 at some point prior to the survey. Among health care workers, only 6% of them reported having been diagnosed of COVID-19 prior to the survey. Additionally, about 37.9% of health care workers reported having at least one chronic condition (results not shown in the table). Table 1 shows a summary of the socio-demographic characteristics of the participants.

**Table 1.** Socio-demographic characteristics of survey participants (N = 1,168)

| Characteristics | n (%) | Vaccine uptake |  |
| --- | --- | --- | --- |
|  |  | Yes (n = 579) | No (n = 589) |
| Age group (years) |  |  |  |
| 18-25 | 125 (10.7) | 53 (42.4) | 72 (57.6) |
| 26-35 | 308 (26.4) | 141 (45.8) | 167 (54.2) |
| 36-45 | 365 (31.3) | 186 (51.0) | 179 (49.0) |
| 46-55 | 214 (18.3) | 109 (50.9) | 105 (49.1) |
| >55 | 156 (13.4) | 90 (57.7) | 66 (42.3) |
| Sex |  |  |  |
| Male | 496 (42.5) | 280 (56.5) | 216 (43.5) |
| Female | 672 (57.5) | 299 (44.5) | 373 (55.5) |
| Health care worker |  |  |  |
| Yes | 420 (36.0) | 234 (55.7) | 186 (44.3) |
| No | 748 (64.0) | 345 (46.1) | 403 (53.9) |
| Chronic conditions |  |  |  |
| Yes | 481 (41.2) | 276 (57.4) | 205 (42.6) |
| No | 687 (58.8) | 303 (44.1) | 384 (55.9) |
| Health status |  |  |  |
| Good | 1054 (90.2) | 520 (49.3) | 534 (50.7) |
| Not so good | 114 (9.8) | 59 (51.8) | 55 (48.2) |
| Tertiary education |  |  |  |
| Yes | 1102 (94.3) | 547 (49.6) | 555 (50.4) |
| No | 66 (5.7) | 32 (48.5) | 34 (51.5) |
| Residence |  |  |  |
| Urban | 1081 (92.6) | 544 (50.3) | 537 (49.7) |
| Rural | 87 (7.4) | 35 (40.2) | 52 (59.8) |
| Employment status |  |  |  |
| Employed | 955 (81.8) | 465 (48.7) | 490 (51.3) |
| Not employed | 103 (8.8) | 58 (56.3) | 45 (43.7) |
| Student | 110 (9.4) | 56 (50.9) | 54 (49.1) |
| Medical Aid |  |  |  |
| Yes | 922 (78.9) | 460 (49.9) | 462 (50.1) |
| No | 246 (21.1) | 119 (48.4) | 127 (51.6) |
| Medical Home |  |  |  |
| Yes | 895 (76.6) | 458 (51.2) | 437 (48.8) |
| No | 273 (23.4) | 121 (44.3) | 152 (57.7) |
| Telehealth access |  |  |  |
| Yes | 665 (56.9) | 341 (51.3) | 324 (48.7) |
| No | 503 (43.1) | 238 (47.3) | 265 (52.7) |
| Got COVID19 |  |  |  |
| Yes | 57 (4.9) | 19 (33.9) | 38 (66.7) |
| No | 1111 (95.1) | 560 (50.4) | 551 (49.6) |

### Uptake of the vaccine

Half (49.9%) of the participants indicated that they would accept the COVID-19 vaccine if they are offered when it becomes available, about 31.1% were unsure of whether they would take the vaccine or not and the other 19% indicated that they would reject the vaccine outright. The proportion of vaccine uptake across the different age groups differed. Among adults aged 55 years and older, about 57.9% were willing to get vaccinated compared to only 43.9% in the 18-25 years age group. Among health care workers, 56.3% reported having an intention to get vaccinated. Additionally, 57.9% of those who reported having at least one chronic condition expressed willingness to get vaccinated once the vaccine was available. Participants with a tertiary education were indifferent about vaccination with 50% expressing interest. After combining responses on our outcome variable, half (49.9%) of the study participants would accept the vaccine, “Yes” (Would Accept) and the other half (50.1%) would not be vaccinated, “No” (Unsure/Would reject).

### Risk of contracting the COVID-19 infection

Among those who had been previously diagnosed of COVID-19, about 37% indicated that their infection was not so severe, while the other 14% did not experience any severe symptoms due to the infection. Additionally, about 37% indicated that they were not worried of the possibility of re-infection by the virus. Approximately 77% of the participants were worried about the possibility that they might contract the virus, while 91% were worried about the possibility that a friend or a relative might catch the virus. Fig 1 shows the perceived risk of contracting COVID-19.

**Fig 1.**
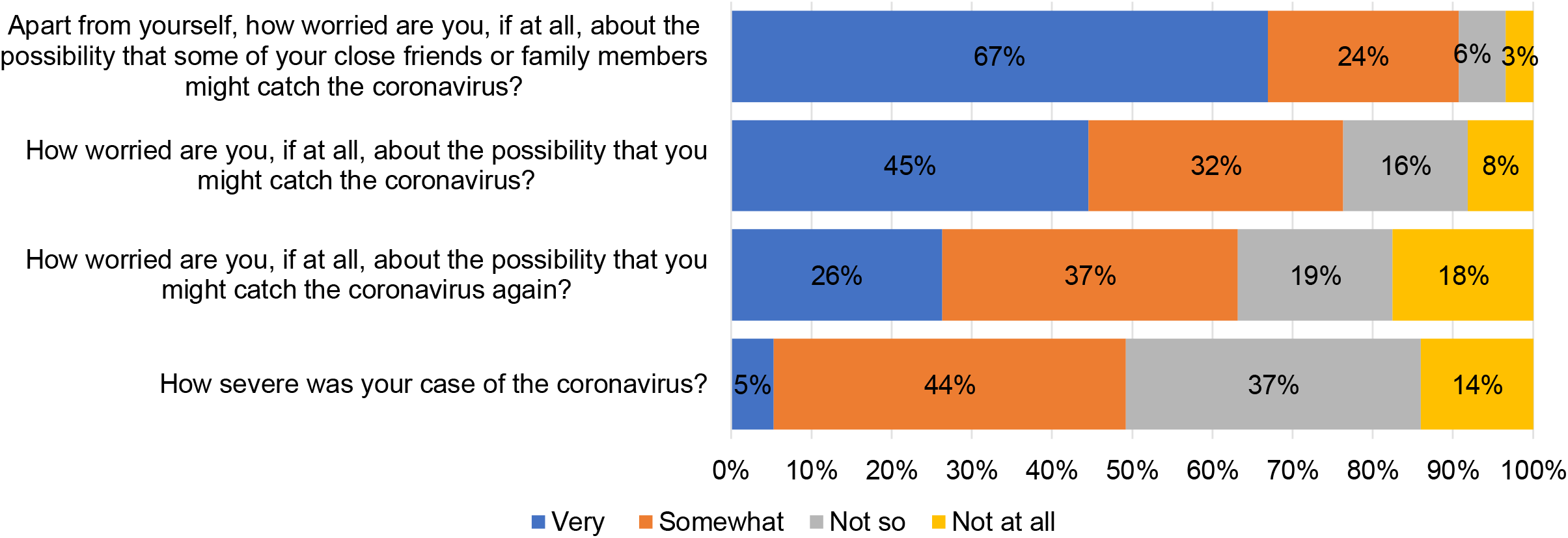
Risk of contracting COVID-19

#### Perceived vaccine effectiveness and safety of the COVID-19 vaccine

Approximately 76% of the participants did not think that the vaccine will be effective in reducing symptoms of the virus and another 72% indicated that they did not think that the vaccine will be effective in preventing infection. Slightly more than half (51%) of the participants indicated that they did not trust the government and other relevant authorities in ensuring that the vaccine is effective and safe. Fig 2 shows the perceived effectiveness and safety of the COVID-19 vaccine.

**Fig 2.**
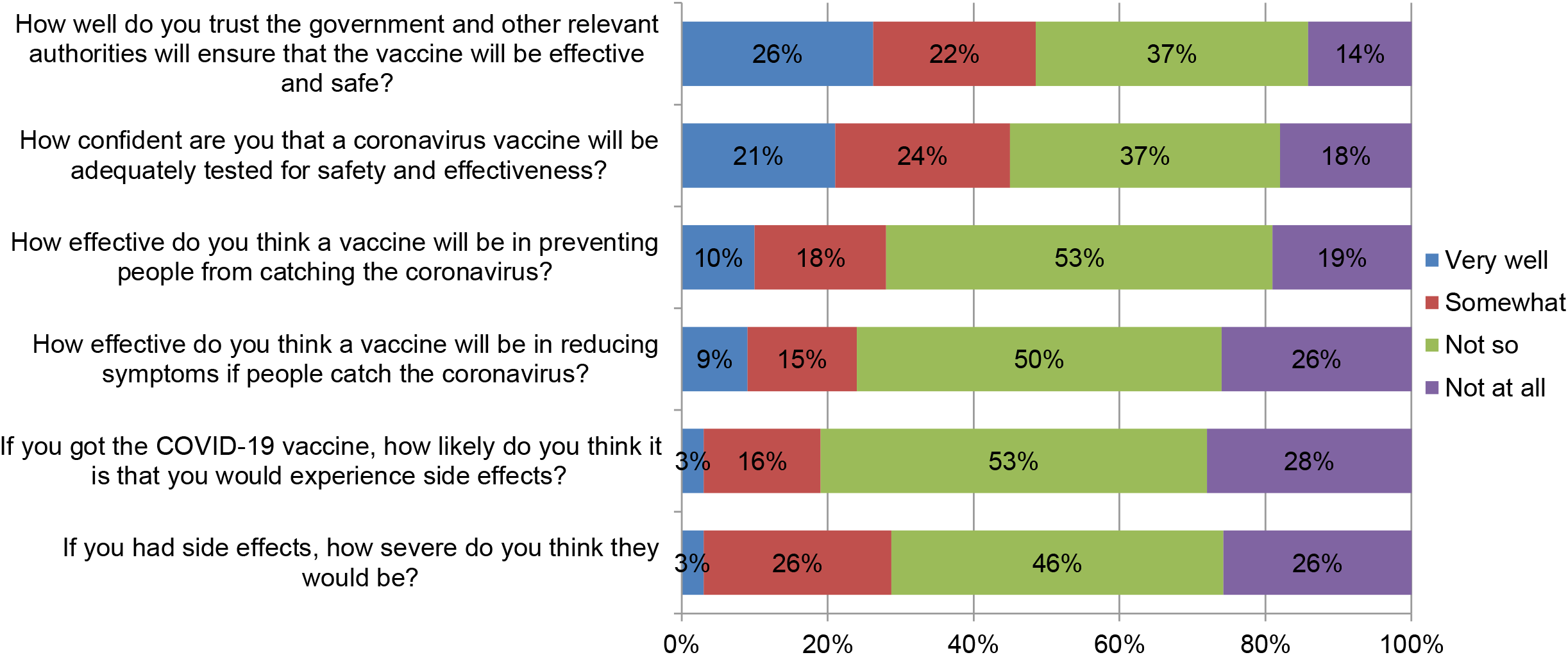
Perceived vaccine effectiveness and safety of the COVID-19 vaccine

#### Decision to be vaccinated with the COVID-19 vaccine

Among factors considered by participants in deciding to get vaccinated or not, we looked at convenience, availability, confidence in vaccine efficacy, vaccine effectiveness, and potential sources of advice. About 58% of the adults indicated that they would strongly consider vaccine effectiveness in their decision-making process. A similar proportion also identified free vaccine access and vaccine safety as important factors in deciding whether to get vaccinated for COVID-19 or not. Fig 3 shows the preferences of the adults on the different factors assessed.

**Fig 3.**
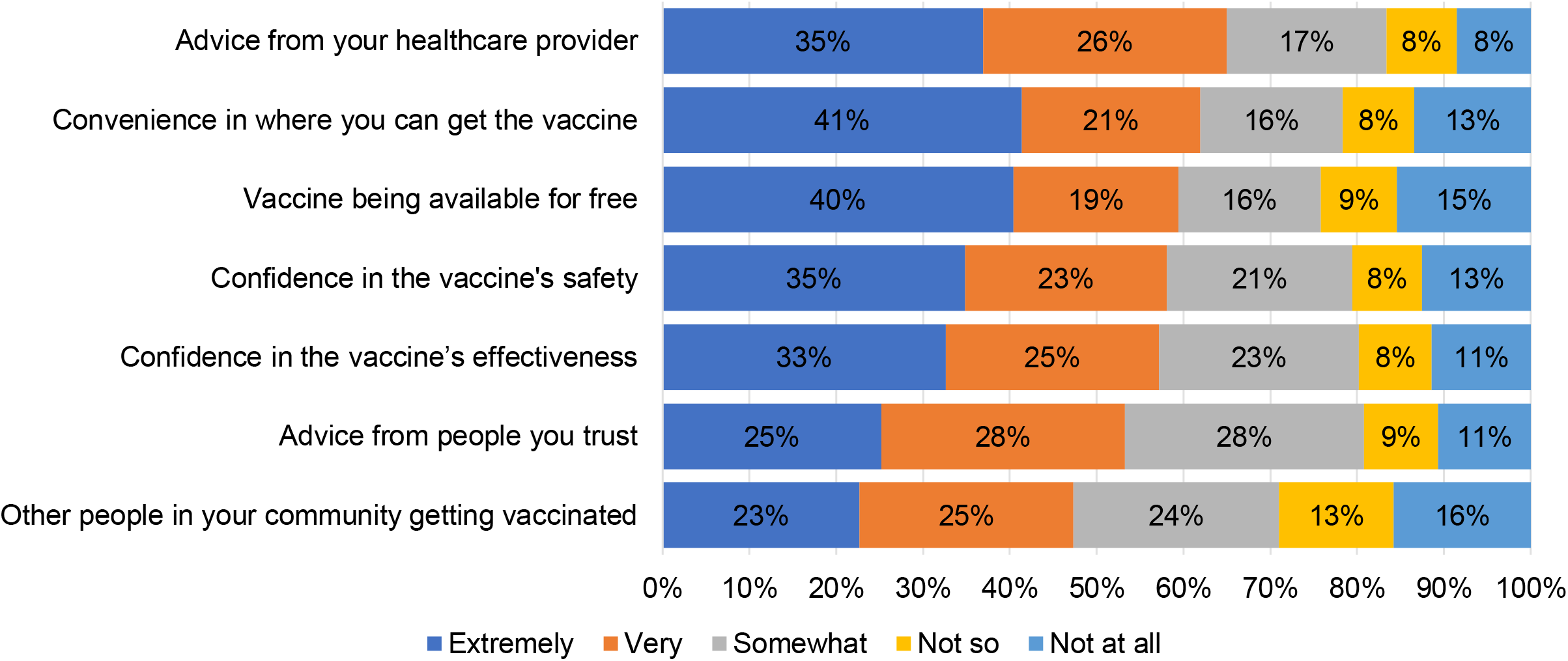
Decision to be vaccinated with the COVID-19 vaccine

#### Factors associated with intention to be vaccinated with the COVID-19 vaccine

Different covariates were assessed on their association with intention to vaccinate using logistic regression analysis (Table 2). After adjusting for all covariates of interests, adults aged 55 years and older were two times more likely to get vaccinated when compared to those aged 18-25 years [Adjusted Odds Ratio (AOR): 2.04, 95% CI: (1.07 – 3.87)]. Similarly, health care workers were 1.73 times more likely to get vaccinated when compared to non-health care workers [AOR: 1.73, [95% CI: (1.34 – 2.24)]. Being male [vs female AOR: 1.84, 95% CI: (1.44 – 2.36)] and having at least one chronic condition [vs no chronic condition AOR: 1.72, 95% CI: (1.32 – 2.25)] were significantly associated with intention to vaccinate. Participants who had been previously diagnosed of COVID-19 [vs no previous COVID-19 infection AOR: 0.45, 95% CI: (0.25 – 0.81)] were significantly less likely to get vaccinated. Not so good health status, tertiary education, and medical home (having a primary health care doctor) were associated with borderline likelihood of getting vaccinated. We assessed the same covariates among the health care workers sub group, and only being male [vs females, AOR: 1.92 95% CI: (1.24 – 2.98)] was associated the likelihood of getting vaccinated.

**Table 2.**
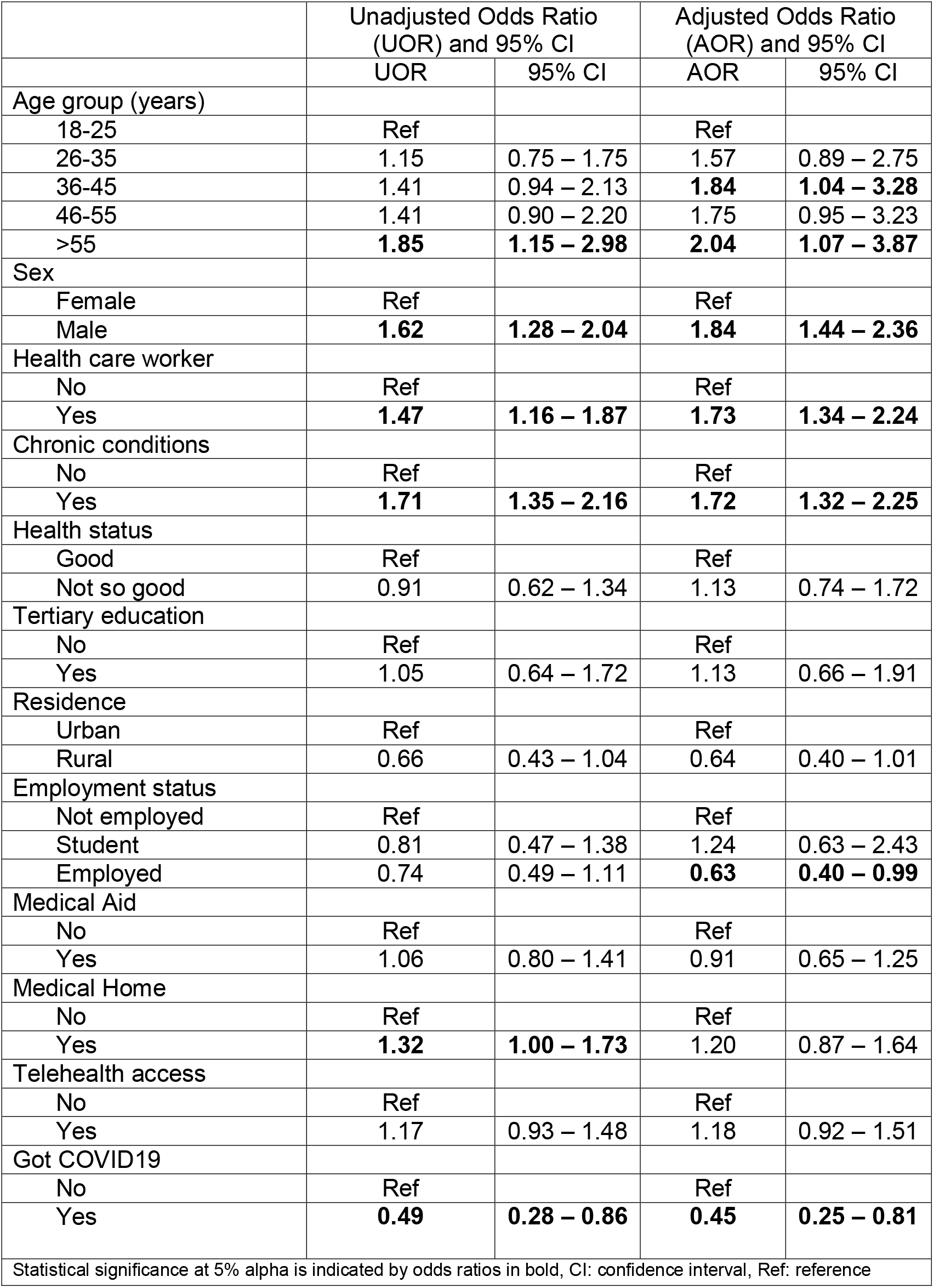
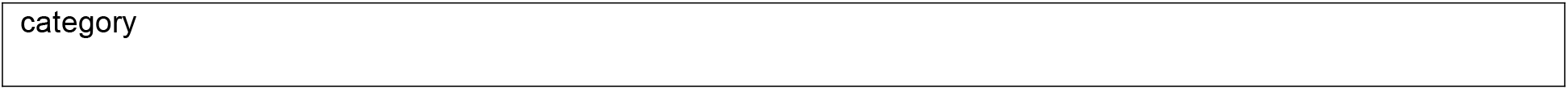
Bivariate and multivariable logistic regression model results

## Discussion

Our study was the first to assess the COVID-19 vaccine hesitancy at national level in Zimbabwe. The study findings showed that half of the study participants reported that they would accept the COVID-19 vaccine if it becomes available. The majority of the survey participants were uncertain about the effectiveness of the vaccine and lacked confidence on the safety of the vaccine. About half lacked trust in the government’s ability to ensure that the vaccine will be effective and most participants would seek the advice of a healthcare worker first before getting vaccinated. Increased age, presence of chronic disease/condition, male gender and being a healthcare worker were associated with increased likelihood of vaccine acceptance while a history of COVID-19 infection and rural residence were associated with reduced likelihood of vaccine acceptance.

Our study findings showed that half of the participants (50%) were willing to take the COVID-19 vaccine while the other half were either unsure or would reject taking the vaccine. These findings are consistent with other African studies from the Democratic Republic of Congo (DRC) (56%) (17), and Nigeria (50%) (18). The similarities in findings may be due to resemblances in the methodology used, as well as socio-economic and political settings in DRC, Nigeria and Zimbabwe. However, a South African survey showed that about 71% were willing to get vaccinated against COVID-19 (19). Based on the significant difference in vaccine acceptance for this study and the South African study, one might hypothesize that South Africans were more likely to accept the vaccine given the high numbers of reported COVID-19 cases and fatalities as compared to the other countries (DRC, Nigeria and Zimbabwe).

Moreover, the surveys from DRC, Nigeria and this study were conducted before the vaccine was rolled out and conspiracy theories on the COVID-19 vaccine were at their peak thus, the researchers predict an increase in vaccine acceptance as more accurate information penetrates the population, more people getting vaccinated and vaccine effectiveness being witnessed in those who would have been vaccinated. The level of vaccine hesitancy found in our study was low when compared to the required COVID-19 herd immunity i.e. 60-70% (20). A COVID-19 vaccine uptake of 50% may not be adequate based on Zimbabwean government’s plans to vaccinate 60% of the population as a way of reaching the herd immunity threshold (21). Therefore, there is a need to come up with strategies to increase the proportion of citizens willing to get the COVID-19 vaccine. Strategies that have been shown to increase COVID-19 vaccine uptake include engagement of community leaders, social mobilization tactics, mass media campaigns, the use of reminder and follow-up systems, training and education of health care professionals, incentives, vaccine mandates, efforts to make vaccine more accessible, and efforts to increase general knowledge and awareness (22,23).

Healthcare workers who took part in this study were more likely to accept the COVID-19 vaccine and this finding corroborated with a study in the U.S (24). It can be hypothesised that healthcare workers perceived their susceptibility to contract COVID-19 as high as evidenced by a study which reported that healthcare workers had a more than seven-fold higher risk of severe COVID-19 across all occupational groups (25). Since health providers have the moral obligation not to harm their patient (26), they may feel compelled to protect the sick and vulnerable patients by getting vaccinated themselves. This is a decisive finding since healthcare workers’ vaccine acceptance can easily untangle vaccine hesitancy when recommending the vaccine to their clients. The health providers can also serve as role models of vaccine acceptance for the general population. Efforts to combat COVID-19 vaccine hesitancy among the general population can utilize the increased intention to vaccinate among health workers by making them community outreach vaccine advocates.

Most of the study participants doubted the effectiveness of the COVID-19 vaccine. This was consistent with findings from an African study where despite the respondents’ agreement with the importance of vaccinating the population against SARS-CoV-2, many had reservations about the effectiveness and safety of the vaccine (27). The doubting of the effectiveness could be attributed to several reasons. Firstly, this could be attributed to the lack of information on how the vaccine was developed and tested. There has been concern on how some vaccines were developed with people preferring those thought to have been developed transparently in comparison to those whose development was shrouded with secrecy (28,29). Secondly, there were concerns on whether trials were not rushed and regulatory standards relaxed considering the unprecedented speed at which the COVID-19 vaccines were developed (30). The lack of lucid information on vaccine development exacerbates the vaccine hesitancy problem and it is essential for health authorities to be equipped with authentic information on how the COVID vaccines were developed; thus enabling them to disseminate factual information and allay the vaccine effectiveness related anxieties.

The perceived lack of acceptance of the COVID-19 vaccine in Zimbabwe can partly be attributed to earlier reports on safety of the COVID-19 vaccines with respect to possible side- and adverse effects (31). Some Zimbabweans became sceptical and reserved towards the vaccine especially after experts questioned the effectiveness of the vaccines developed before the 501.V2 coronavirus variant which originated from South Africa and appeared to be a more contagious strain infiltrating the country (21). The widespread dissemination of vaccine safety information should be done before rolling out the vaccine to ensure that people make an informed decision based on scientifically proven information instead of rumours and conspiracy theories. The health authorities should continue to update citizens about vaccine side-effects and institute strong pharmacovigilant systems as well as, compensation schemes for severe adverse events as this might boost public confidence in vaccine safety (32,33).

About half of the participants in this study lacked trust on whether the government and relevant authorities would be able to provide a safe vaccine that would protect them from COVID-19. Such public insecurities were also observed in other studies on vaccine hesitancy (34,35). These public insecurities may be caused by unsubstantiated political statements, incapacities of the health system and centralisation of health services. Unscientific claims on COVID-19 by individuals in positions of authority can introduce uncertainties within the population and lower their trust in how the government is handling the pandemic. The Zimbabwean healthcare system is facing serious challenges including shortage of essential drugs, lack of equipment to carry out basic procedures, and skills migration thus, citizens may lack trust in the system’s ability to contain the pandemic. Despite the proposed free COVID-19 vaccination, there are other indirect costs that could be incurred by citizens to get to the vaccination sites which are currently centralised at secondary and tertiary health facilities. Travelling costs and loss of productive time could be potential barriers to COVID-19 vaccination in low-income settings like Zimbabwe. Thus, the government should work towards increasing the national health budget to cater for the excess service demand caused by the pandemic. It is also critical to decentralize vaccination sites and to use influential community leaders for community outreach activities.

The current study also revealed that an increase in age and the presence of a chronic condition were associated with a high likelihood of vaccine acceptance. These findings are consistent with findings from other studies that showed a strong association between age and willingness to get vaccinated (35,36). The risk of severe COVID-19 increases with age and 80% of COVID-19 deaths are of adults 65 years and older (37). Additionally, multiple chronic conditions complicate the progression of COVID-19 (38). Thus, the elderly and those with chronic conditions may have increased perceived vulnerability to the disease and are more likely to take steps towards protecting themselves.

We noted that males were more likely to accept the vaccine. Several other studies also revealed similar results (27,28,39–41). This was an unexpected finding given the poor health seeking behaviour among men (42). However, men constitutes the highest proportion of the Zimbabwean workforce and the speculations that some organisations might prioritise or mandate vaccination among their employees to continue productivity in wake of anticipated lockdowns might have influenced men’s decision. The reduced vaccine acceptance among women can be attributed to a lack of information of the safety of the vaccine especially with regards to conception related matters; however experts believe that the vaccines were unlikely to pose a risk to pregnant and lactating woman (43). The finding that among the health care worker subgroup, males were also likely to be vaccinated than females can be explained by some of the reasons discussed above. Health authorities have the responsibility to give women updated information on the safety of the vaccine during the community outreach activities. It was surprising to note that 39% of those who reported previous COVID-19 infection in this study were less likely to be vaccinated. This is despite the fact that those with a history of prior infection could be reinfected by SARS-CoV-2 even after recovering from the initial infection (44). Studies have shown that primary infection only provides short term protective immunity against SARS-CoV-2 (45–47). Individuals who would have recovered from coronavirus infection may perceive their susceptibility as low assuming that they would have garnered natural immunity. Although those who have been previously infected by SARS-CoV-2 are less likely to have a repeat infection when compared to those without evidence of previous infection, they are still susceptible to reinfection (48), hence, the need to be vaccinated as well.

The present study noted that rural residents were less likely to pursue COVID-19 vaccination when compared to those residing in urban areas. This finding corroborates with findings from other similar studies (28,34,49). This could be attributed to poor information penetration and reduced perceived susceptibility risk among the rural folks. The finding can also be explained by the fact that rural residents have reduced health literacy and awareness, reduced trust and interaction with healthcare workers, and presumed cost-based concerns (34,50,51). Information travels faster in urban areas where mass media and internet connection are readily available thus, it can be easier for those residing in urban areas to access authentic information about the vaccine when compared to most remote rural areas in Zimbabwe. This shows the importance of tailoring vaccine awareness strategies relative to populations residing in varying geographical regions. The authors recommend that policy makers consider geographical- and socio-cultural-specific information for education and communication when approaching the diverse residential settings.

## Strengths and limitations

Our study assessed a broad range of factors that have been known to influence vaccine acceptance. The findings can be used to guide future health activities with an aim of improving COVID-19 vaccine uptake or uptake of other vaccines. All the ten Zimbabwean provinces were represented although some were more represented than others. The data collection tools were distributed in all the three common languages used in Zimbabwe to accommodate the majority of citizens. The study received a fairly good response rate, which was unexpected by the researchers given that no incentives were given to cover internet connection fees. The increased use of internet services during the COVID-19 era enabled swift and cost effective online data collection on the part of the researchers.

Despite the relatively large sample size, the generalizability of our findings can be hampered by our sampling method. The researchers used mainly their social media networks as proxies for distributing the data collection tool. The study was also limited to those who had access to mobile phones, tablets, or computers, thus introducing a selection bias. We may have excluded the poor and the old who in fact are vulnerable to COVID-19. Besides, the data were collected before the vaccine was rolled out in the country and vaccine hesitancy may have waned as more authentic information became available to the population. The participants were mostly urban dwellers with easy access to internet connection and this could have potentially resulted in a selection bias leaving the rural population. The fact that this study utilized self-reported data makes it vulnerable to reporting bias.

There is a need for further studies to understand how COVID-19 vaccine hesitancy will evolve with time. With the COVID-19 vaccine now available in the country and already being given, studies should be conducted periodically to assess the time-sensitive aspect of the vaccine hesitancy among those who rejected the vaccine or were unsure of it. Our study reported the quantitative findings associated with COVID-19 vaccine; there is therefore a need for qualitative studies to explore contextual and other individual factors that lead to vaccine hesitancy. Since in our study we were unable to report vaccine hesitancy among targeted sectors of the populations e.g. education, commerce, tourism among others, studies are required to assess how vaccine hesitancy will vary across these sectors. These will enable targeted messaging to improve the vaccine uptake. Though we assessed vaccine hesitancy among health care workers, we were unable to disentangle our data to find out hesitancy among various groups of healthcare workers. Future studies looking into vaccine hesitancy among health care workers should be in position to assess vaccine hesitancy among doctors, nurses, pharmacists and many other healthcare worker professionals. To try and be more representative, especially inclusion of the rural communities, future studies should be conducted physically so that the barrier of access to internet and electronic gadgets can be bridged. However, strategies to mitigate against COVID-19 infection and spread should be observed.

## Conclusions

In our study, we found half of the participants were willing to get vaccinated against COVID-19 with the majority lacking trust in the government and being uncertain about vaccine effectiveness and safety. The high vaccine acceptance among health workers and the finding that the majority will consult health care workers before deciding to vaccinate, can be a good foundation to launch a successful COVID-19 vaccine awareness campaign in the country. However, the level of vaccination uptake is way below the expected herd immunity hence the government and other relevant authorities should provide timely and accurate information through community-wide campaigns. Strategies should also be put in place to target and prioritise groups likely not to vaccinate such as females, younger ages and those without chronic conditions. The government and other relevant authorities should aim for a more transparent strategy since it will contribute to public trust and increased acceptability of the vaccine.

## Data Availability

Data used during analysis is available from the corresponding author upon a reasonable request.

## Acknowledgements

The authors would like to thank the following organisations for the support during the study; the University of Zimbabwe, Faculty of Medicine and Health Sciences, Global and Public Health Unit, Africa University, College of Health, Agriculture & Natural Science, Clinical Research Centre (AUCRC) and the Zimbabwe College of Public Health Physicians (ZCPHP). The authors would like to thank the following individuals Mandla Tirivavi from AUCRC for administrative support, Brian Maponga and Pamela Magande from the ZCPHP for facilitating the distribution of the survey and ethical approval. Finally, we thank all the Zimbabweans who participated in the survey.

## Authors Contributions

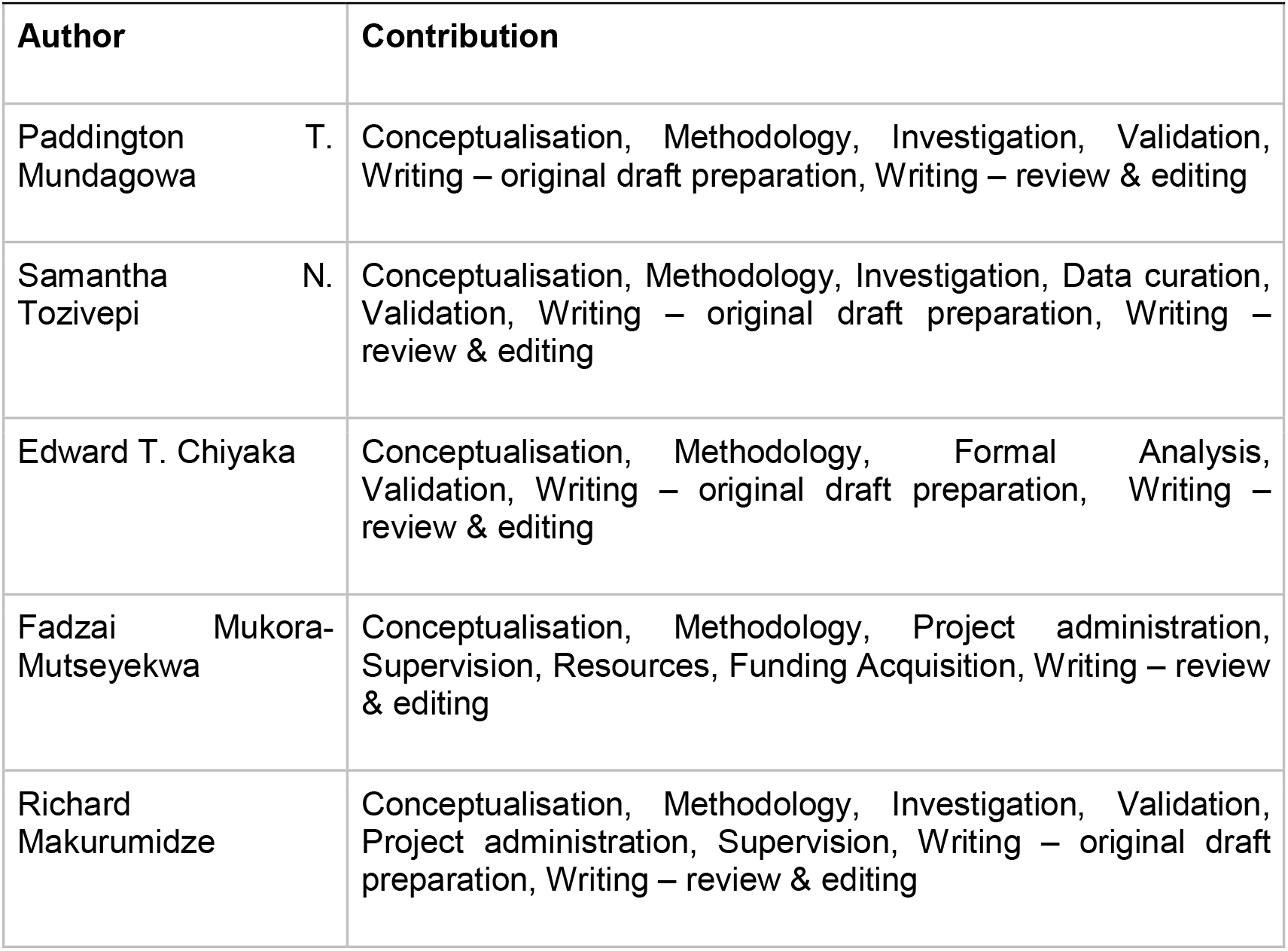

## Declarations

### Data Availability

The data used during analysis is available from the corresponding author upon a reasonable request.

## References

1. WHO. WHO Coronavirus (COVID-19) Dashboard [Internet]. 2021 [cited 2021 May 28]. Available from: https://covid19.who.int/

2. Jahanshahi AA, Dinani MM, Madavani AN, Li J, Zhang SX. The distress of Iranian adults during the Covid-19 pandemic – More distressed than the Chinese and with different predictors. Brain Behav Immun [Internet]. 2020 Jul;87:124–5. Available from: https://linkinghub.elsevier.com/retrieve/pii/S0889159120307121

3. Mukaetova-Ladinska EB, Kronenberg G. Psychological and neuropsychiatric implications of COVID-19. Eur Arch Psychiatry Clin Neurosci [Internet]. 2021 Mar 22;271(2):235–48. Available from: http://link.springer.com/10.1007/s00406-020-01210-2

4. Morgantini LA, Naha U, Wang H, Francavilla S, Acar Ö, Flores JM, Crivellaro S, Moreira D, Abern M, Eklund M, Vigneswaran HT WS. Factors contributing to healthcare proffessional burnout during the COVID-19 Pandemic: A rapid turnaround global survey. PLoS One. 2020;15(9).

5. WHO. Draft Landscape of COVID-19 Candidate Vaccines [Internet]. Geneva; 2021. Available from: https://www.who.int/publications/m/item/draft-landscape-of-covid-19-candidate-vaccines

6. Anderson RM, Vegvari C, Truscott J, Collyer BS. Challenges in creating herd immunity to SARS-CoV-2 infection by mass vaccination. Lancet [Internet]. 2020 Nov;396(10263):1614–6. Available from: https://linkinghub.elsevier.com/retrieve/pii/S0140673620323187

7. WHO. Ten threats to global health in 2019 [Internet]. World Health Organization. 2019 [cited 2021 May 28]. Available from: https://www.who.int/news-room/spotlight/ten-threats-to-global-health-in-2019

8. WHO. Infodemic management-infodermiology [Internet]. World Health Organization. 2020 [cited 2021 Jan 19]. Available from: https://www.who.int/teams/risk-communication/infodemic-msnsgement

9. MacDonald NE. Vaccine hesitancy: Definition, scope and determinants. Vaccine [Internet]. 2015 Aug;33(34):4161–4. Available from: https://linkinghub.elsevier.com/retrieve/pii/S0264410X15005009

10. MoHCC. COVID-19 vaccination gets traction [Internet]. Ministry of Health and Child Care, Zimbabwe. 2021 [cited 2021 Jan 19]. Available from: http://www.mohcc.gov.zw/index.php?option=com_content&view=article&id=355:covid-19-vaccination-gets-traction&catid=84&Itemid=435

11. Worldometer. Zimbabwe population 2021 [Internet]. Worldometer. 2021 [cited 2021 May 28]. Available from: https://www.worldometers.info/world-population/zimbabwe-population/#:∼:text=Zimbabwe 2020 population is estimated,year according to UN data.

12. ZIMSTAT. Census 2012 Preliminary Report. Harare; 2012.

13. Ministry of Health and Child Care(MoHCC). Situation report COVID-19, Zimbabwe. Harare; 2021.

14. Topline Research Solutions. Zimbabweans opinion survey: How best to assist the COVID 19 situation in Zimbabwe [Internet]. Harare; 2020. Available from: https://www.google.com/url?sa=t&rct=j&q=&esrc=s&source=web&cd=&ved=2ahUKEwj5ieON1fTwAhWFGuwKHeoCBWIQFjABegQIAxAD&url=https%3A%2F%2Ftopliners.co.za%2Fwp-content%2Fuploads%2F2021%2F01%2FProject-Hondo-FINAL-Report-14-Jan-2021.pdf&usg=AOvVaw2pLvgfxJPnqKi4xAlg

15. Lin C, Tu P, Beitsch LM. Confidence and receptivity for covid-19 vaccines: A rapid systematic review. Vaccines. 2021;9(1):1–32.

16. COVID Collaborative. Coronavirus Vaccine Hesitancy in Black and Latinx Communities [Internet]. COVID Collaborative. 2020 [cited 2021 Jun 6]. Available from: https://www.covidcollaborative.us/content/vaccine-treatments/coronavirus-vaccine-hesitancy-in-black-and-latinx-communities

17. Ditekemena JD, Nkamba DM, Mutwadi A, Mavoko HM, Siewe Fodjo JN, Luhata C, et al. COVID-19 Vaccine Acceptance in the Democratic Republic of Congo: A Cross-Sectional Survey. Vaccines [Internet]. 2021 Feb 14;9(2):153. Available from: https://www.mdpi.com/2076-393X/9/2/153

18. Tobin E, Okonofua M, Azeke A, Ajekweneh V, Akpede G. Willingness to acceptance a COVID-19 vaccine in Nigeria: a population-based cross-sectional study. J Med Res. 2021;5(2):1–6.

19. Monama T. 71% of South Africans are willing to take the COVID-19 vaccine [Internet]. 2021. Available from: https://www.news24.com/news24/southafrica/news/71-of-south-africans-are-willing-to-take-the-covid-19-vaccine-study-20210512

20. Wang W, Wu Q, Yang J, Dong K, Chen X, Bai X, et al. Global, regional, and national estimates of target population sizes for covid-19 vaccination: descriptive study. BMJ [Internet]. 2020 Dec 15;m4704. Available from: https://www.bmj.com/lookup/doi/10.1136/bmj.m4704

21. Mavhunga C. Government urges Zimbabweans to accept COVID-19 Vaccine. VOA News [Internet]. 2021 Feb 13; Available from: https://www.voanews.com/covid-19-pandemic/government-urges-zimbabweans-accept-covid-19-vaccine

22. Jarrett C, Wilson R, O’Leary M, Eckersberger E, Larson HJ. Strategies for addressing vaccine hesitancy – A systematic review. Vaccine [Internet]. 2015 Aug;33(34):4180–90. Available from: https://linkinghub.elsevier.com/retrieve/pii/S0264410X15005046

23. Gayle H, Foege W, Brown L, Kahn B. Framework for Equitable Allocation of COVID-19 Vaccine: Achieving Acceptance of COVID-19 Vaccine. National Academies of Sciences, Engineering, and Medicine. 2020.

24. Shaw J, Stewart T, Anderson KB, Hanley S, Thomas SJ, Salmon DA, et al. Assessment of US Healthcare Personnel Attitudes Towards Coronavirus Disease 2019 (COVID-19) Vaccination in a Large University Healthcare System. Clin Infect Dis [Internet]. 2021 Jan 25; Available from: https://academic.oup.com/cid/advance-article/doi/10.1093/cid/ciab054/6118651

25. Mutambudzi M, Niedzwiedz C, Macdonald EB, Leyland A, Mair F, Anderson J, et al. Occupation and risk of severe COVID-19: prospective cohort study of 120 075 UK Biobank participants. Occup Environ Med [Internet]. 2021 May;78(5):307–14. Available from: https://oem.bmj.com/lookup/doi/10.1136/oemed-2020-106731

26. Gur-Arie R, Jamrozik E, Kingori P. No Jab, No Job? Ethical Issues in Mandatory COVID-19 Vaccination of Healthcare Personnel. BMJ Glob Heal [Internet]. 2021 Feb 17;6(2):e004877. Available from: https://gh.bmj.com/lookup/doi/10.1136/bmjgh-2020-004877

27. Africa Centres for Disease Control and Prevention, London School of Hygiene and Tropical Medicine’s Vaccine Confidence Project, Orb International. COVID 19 Vaccine Perceptions: A 15 country study. 2021;(February):1–70.

28. El-Elimat T, AbuAlSamen MM, Almomani BA, Al-Sawalha NA, Alali FQ. Acceptance and attitudes toward COVID-19 vaccines: A cross-sectional study from Jordan. medRxiv [Internet]. 2020;(816):1–15. Available from: http://dx.doi.org/10.1371/journal.pone.0250555

29. Pogue K, Jensen JL, Stancil CK, Ferguson DG, Hughes SJ, Mello EJ, et al. Influences on Attitudes Regarding Potential COVID-19 Vaccination in the United States. Vaccines [Internet]. 2020 Oct 3;8(4). Available from: http://www.ncbi.nlm.nih.gov/pubmed/33022917

30. Shah A, Marks PW, Hahn SM. Unwavering Regulatory Safeguards for COVID-19 Vaccines. JAMA [Internet]. 2020 Sep 8;324(10):931. Available from: https://jamanetwork.com/journals/jama/fullarticle/2769421

31. Everington K. Full list of adverse reactions from China’s Sinopharm vaccine revealed. Taiwan News [Internet]. 2021 Jan 11; Available from: https://www.taiwannews.com.tw/en/news/4098913

32. Chandler RE. Optimizing safety surveillance for COVID-19 vaccines. Nat Rev Immunol [Internet]. 2020 Aug 17;20(8):451–2. Available from: http://www.nature.com/articles/s41577-020-0372-8

33. Halabi S, Heinrich A, Omer SB. No-Fault Compensation for Vaccine Injury — The Other Side of Equitable Access to Covid-19 Vaccines. N Engl J Med [Internet]. 2020 Dec 3;383(23):e125. Available from: http://www.nejm.org/doi/10.1056/NEJMp2030600

34. Fisher KA, Bloomstone SJ, Walder J, Crawford S, Fouayzi H, Mazor KM. Attitudes Toward a Potential SARS-CoV-2 Vaccine. Ann Intern Med [Internet]. 2020 Dec 15;173(12):964–73. Available from: https://www.acpjournals.org/doi/10.7326/M20-3569

35. Soares P, Rocha JV, Moniz M, Gama A, Laires PA, Pedro AR, et al. Factors associated with COVID-19 vaccine hesitancy. Vaccines. 2021;9(3):1–14.

36. Lazarus J V., Wyka K, Rauh L, Rabin K, Ratzan S, Gostin LO, et al. Hesitant or Not? The Association of Age, Gender, and Education with Potential Acceptance of a COVID-19 Vaccine: A Country-level Analysis. J Health Commun [Internet]. 2020 Oct 2;25(10):799–807. Available from: https://www.tandfonline.com/doi/full/10.1080/10810730.2020.1868630

37. CDC. Older Adults at greater risk of requiring hospitalization or dying if diagnosed with COVID-19 [Internet]. Center for Disease Control and Prevention. 2021. Available from: https://www.cdc.gov/coronavirus/2019-ncov/need-extra-precautions/older-adults.html

38. Liu K, Chen Y, Lin R, Han K. Clinical features of COVID-19 in elderly patients: A comparison with young and middle-aged patients. J Infect [Internet]. 2020 Jun;80(6):e14.#x2013;8. Available from: https://linkinghub.elsevier.com/retrieve/pii/S016344532030116X

39. Alqudeimat Y, Alenezi D, AlHajri B, Alfouzan H, Almokhaizeem Z, Altamimi S, et al. Acceptance of a COVID-19 Vaccine and its Related Determinants among the General Adult Population in Kuwait. Med Princ Pract [Internet]. 2021 Jan 22; Available from: https://www.karger.com/Article/FullText/514636

40. Malik AA, McFadden SM, Elharake J, Omer SB. Determinants of COVID-19 vaccine acceptance in the US. EClinicalMedicine [Internet]. 2020 Sep;26:100495. Available from: https://linkinghub.elsevier.com/retrieve/pii/S258953702030239X

41. Wong LP, Alias H, Wong P-F, Lee HY, AbuBakar S. The use of the health belief model to assess predictors of intent to receive the COVID-19 vaccine and willingness to pay. Hum Vaccin Immunother [Internet]. 2020 Sep 1;16(9):2204–14. Available from: https://www.tandfonline.com/doi/full/10.1080/21645515.2020.1790279

42. Mufunda E, Albin B, Hjelm K. Differences in Health and Illness Beliefs in Zimbabwean Men and Women with Diabetes. Open Nurs J [Internet]. 2012 Aug 6;6:117–25. Available from: http://benthamopen.com/ABSTRACT/TONURSJ-6-117

43. CDC. COVID-19 Vaccines While Pregnant or Breastfeeding [Internet]. Center for Disease Control and Prevention. 2021 [cited 2021 May 28]. Available from: https://www.cdc.gov/coronavirus/2019-ncov/vaccines/recommendations/pregnancy.html

44. Tillett RL, Sevinsky JR, Hartley PD, Kerwin H, Crawford N, Gorzalski A, et al. Genomic evidence for reinfection with SARS-CoV-2□: a case study. Lancet Infect Dis [Internet]. 2021;21(1):52–8. Available from: http://dx.doi.org/10.1016/S1473-3099(20)30764-7

45. Abu-Raddad LJ, Chemaitelly H, Malek JA, Ahmed AA, Mohamoud YA, Younuskunju S, et al. Assessment of the Risk of Severe Acute Respiratory Syndrome Coronavirus 2 (SARS-CoV-2) Reinfection in an Intense Reexposure Setting. Clin Infect Dis. 2020;2.

46. Edridge E, Kaczorowska J, Hoste A, Baker M, Klein M, Loens K, et al. Seasonal coronavirus protective immunity is short-lasting. Nat Med. 2020;11:1691–3.

47. McMahon A, Robb NC. Reinfection with SARS-CoV-2: Discrete SIR (Susceptible, Infected, Recovered) Modeling Using Empirical Infection Data. JMIR Public Heal Surveill [Internet]. 2020 Nov 16;6(4):e21168. Available from: http://publichealth.jmir.org/2020/4/e21168/

48. Letizia AG, Ge Y, Vangeti S, Goforth C, Weir DL, Kuzmina NA, et al. Articles SARS-CoV-2 seropositivity and subsequent infection risk in healthy young adults□: a prospective cohort study. Lancet Respir [Internet]. 2021;2600(21):1–9. Available from: http://dx.doi.org/10.1016/S2213-2600(21)00158-2

49. Khubchandani J, Sharma S, Price JH, Wiblishauser MJ, Sharma M, Webb FJ. COVID-19 Vaccination Hesitancy in the United States: A Rapid National Assessment. J Community Health [Internet]. 2021 Apr 3;46(2):270–7. Available from: http://link.springer.com/10.1007/s10900-020-00958-x

50. Ferdinand KC, Nedunchezhian S, Reddy TK. The COVID-19 and Influenza “Twindemic”: Barriers to Influenza Vaccination and Potential Acceptance of SARS-CoV2 Vaccination in African Americans. J Natl Med Assoc [Internet]. 2020 Dec;112(6):681–7. Available from: https://linkinghub.elsevier.com/retrieve/pii/S0027968420304089

51. Quinn SC, Jamison AM, Freimuth V. Communicating Effectively About Emergency Use Authorization and Vaccines in the COVID-19 Pandemic. Am J Public Health [Internet]. 2021 Mar;111(3):355–8. Available from: https://ajph.aphapublications.org/doi/full/10.2105/AJPH.2020.306036

